# Preparedness and vulnerability of African countries against introductions of 2019-nCoV

**DOI:** 10.1101/2020.02.05.20020792

**Authors:** Marius Gilbert, Giulia Pullano, Francesco Pinotti, Eugenio Valdano, Chiara Poletto, Pierre-Yves Boëlle, Eric D’Ortenzio, Yazdan Yazdanpanah, Serge Paul Eholie, Mathias Altmann, Bernardo Gutierrez, Moritz U.G. Kraemer, Vittoria Colizza

## Abstract

**Background:** The novel coronavirus (2019-nCoV) epidemic has spread to 23 countries from China. Local cycles of transmission already occurred in 7 countries following case importation. No African country has reported cases yet. The management and control of 2019-nCoV introductions heavily relies on country’s health capacity. Here we evaluate the preparedness and vulnerability of African countries against their risk of importation of 2019-nCoV.

**Methods:** We used data on air travel volumes departing from airports in the infected provinces in China and directed to Africa to estimate the risk of introduction per country. We determined the country’s capacity to detect and respond to cases with two indicators: preparedness, using the WHO International Health Regulation Monitoring and Evaluation Framework; and vulnerability, with the Infectious Disease Vulnerability Index. Countries were clustered according to the Chinese regions contributing the most to their risk.

**Findings:** Countries at the highest importation risk (Egypt, Algeria, Republic of South Africa) have moderate to high capacity to respond to outbreaks. Countries at moderate risk (Nigeria, Ethiopia, Sudan, Angola, Tanzania, Ghana, Kenya) have variable capacity and high vulnerability. Three clusters of countries are identified that share the same exposure to the risk originating from the provinces of Guangdong, Fujian, and Beijing, respectively.

**Interpretation:** Several countries in Africa are stepping up their preparedness to detect and cope with 2019-nCoV importations. Resources and intensified surveillance and capacity capacity should be urgently prioritized towards countries at moderate risk that may be ill-prepared to face the importation and to limit onward transmission.

**Funding:** This study was partially supported by the ANR project DATAREDUX (ANR-19-CE46-0008-03) to VC; the EU grant MOOD (H2020-874850) to MG, CP, MK, PYB, VC.

## Introduction

On January 30, 2020, the World Health Organization (WHO) declared the current novel coronavirus (2019-nCoV) epidemic a Public Health Emergency of International Concern [1]. As of February 1, the epidemic registered 11,818 cases in China and spread to 23 countries that reported a total of 127 cases [2]. Limited local transmission outside China has been reported in Germany, France, Japan, South Korea, Thailand, Vietnam, and the United States.

All continents reported confirmed cases of 2019-nCoV, except Africa. China, however, is the leading commercial partner for Africa, and this generates large travel volumes through which the novel coronavirus might eventually reach the continent. Several measures have already been put in place to prevent and control possible case importations from China [3,4].

The ability to limit and control local transmission following introduction depends, however, on the application and execution of strict measures of detection, prevention and control. These include heightened surveillance, rapid identification of suspect cases followed by patient transfer and isolation, rapid diagnosis, tracing and follow-up of potential contacts [1]. The application of such a vast technical and operational set of interventions depends on countries’ public health and laboratory infrastructures and resources.

Here we assessed the risk of importation to Africa of 2019-nCoV cases from affected provinces in China and put it in relation with countries’ vulnerability to epidemic emergencies and capacity to respond. Importation risk is determined by the volumes of the air traffic connections [5-9] from areas where the virus currently circulates in China. Countries’ functional capacity to manage health security issues is based on WHO International Health Regulation (IHR) Monitoring and Evaluation Framework (MEF) [10], and on an indicator of vulnerability to emerging epidemics.

## Methods

The risk of importation of 2019-nCoV cases to Africa from China was estimated based on origin-destination air-travel flows from January 2019 [8,11,12], number of cases in Chinese provinces, and the population in each of the Chinese provinces that report transmission. Case data were compiled as part of an international effort to build a database at individual level, and included all confirmed cases recorded until the 27th January 2020 [13]. Human population data per province [14] was used to estimate incidence in China. Province-level incidence data were linked to the three airports with the largest volumes in each province [12] (Fig. 1). The province of Hubei was not included among the possible locations that can export the virus, given the travel ban put in place by Chinese authorities on January 23rd and 24th [5]. The importation risk per country in Africa measured the probability of importing a case from the infected provinces in China, accounting for the origin-destination travel flows originated from such provinces and for their different epidemic levels (see appendix for details).

**Figure 1.**
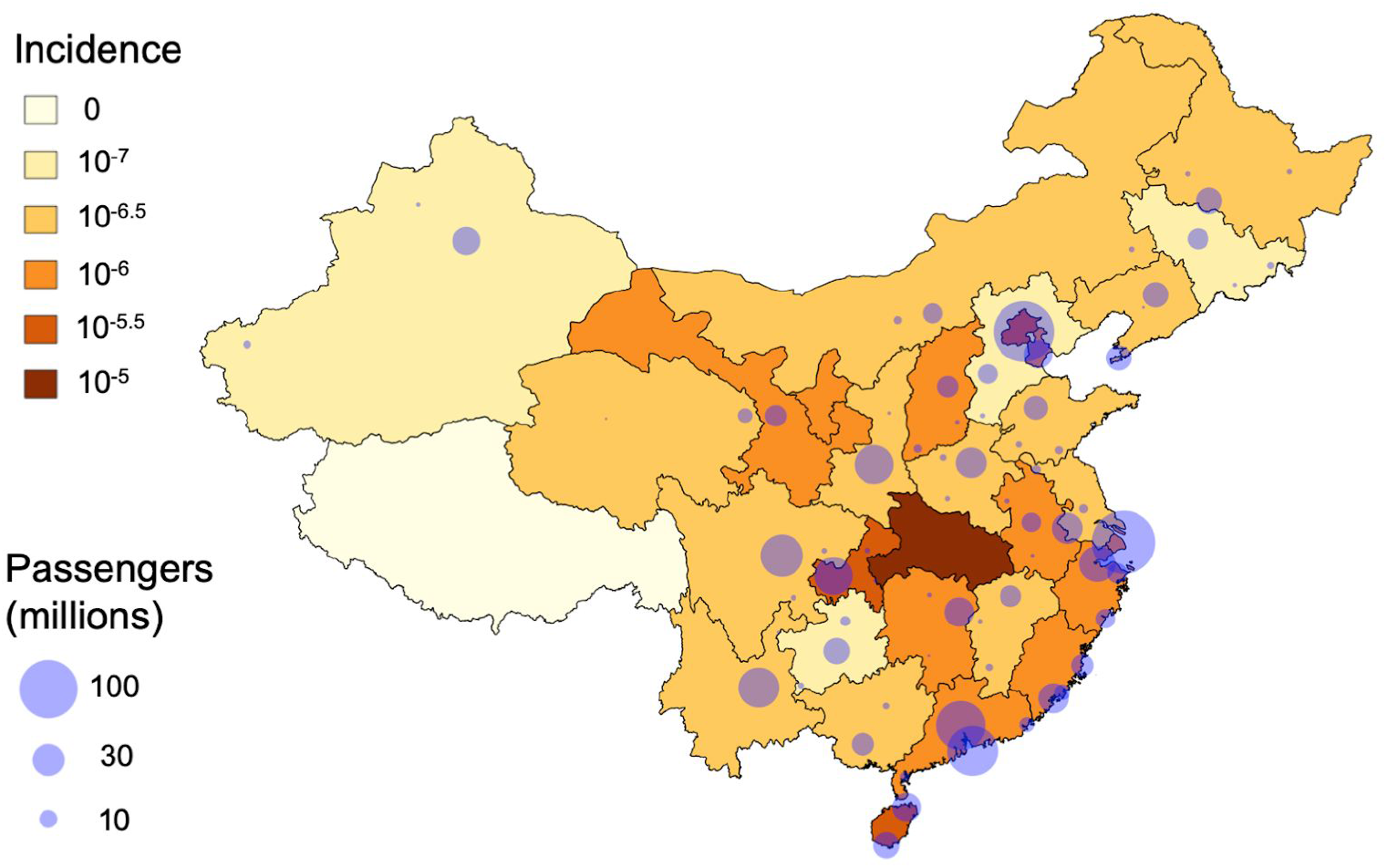
2019-nCoV incidence in China as of 27th January 2020 [13] and annual volume of outflow passenger per airport [12].

For each African country, the most likely origins of the potential case importation were identified. This was done by computing a country’s exposure to each Chinese province, measuring the probability for a city in China to be the origin of a travelling case to the country. Similarity between exposure profiles of different countries was quantified with entropy-based metrics [15], and used to group countries with similar importation patterns via agglomerative clustering (see appendix for details).

Indicators of capacity were taken from the WHO International Health Regulation (IHR) Monitoring and Evaluation Framework (MEF), a set of four components developed by WHO in consultation with partners to support the evaluation of countries’ functional ability to manage health security issues. The MEF is composed of a mandatory self-reporting of capacity (the State Parties self-assessment Annual Reporting, SPAR [10]), and three voluntary components, namely the Joint External Evaluation (JEE), the after-action reviews (AAR) and simulation exercises (SimEx) (all collected and disseminated by WHO).

The 2018 SPAR database [16] contained a total of 20 indicator scores on a scale of 0 to 100, organized and grouped according to the following capacities (number between brackets is the number of indicators per capacity, see [10]): Legislation (2), IHR Coordination (2), Zoonoses (1), Food safety (1), Laboratory (3), Surveillance (2), Human resource (1), National health emergency framework (3), Health service provision (3), Communication (1), Points of Entry (2), Chemical events (1) and Radiation emergency (1). From them, we derived two aggregated indicators that would be relevant to quantify the countries capacity to deal with the introduction and spread of 2019-nCoV. The first quantifies the overall capacity of a country to deal with an emergency linked to a directly transmitted infectious disease, from early detection to the overall handling of a potential epidemic, and was estimated as an average between all indicators, except those of the groups Zoonoses, Food safety, Chemical events, Radiation emergency. The second was more focused on early detection, and averaged the scores of the groups Laboratory, Surveillance, Points of Entry. A very high correlation between the two metrics was found (All countries: *r* = 0.929, *n* = 182, *p* < 0.001; Africa : *r* = 0.893, *n* = 52, *p* < 0.001), so we considered only the first metric that we call SPAR indicator in the following analyses.

The vulnerability of countries to epidemic risk is measured with the Infectious Disease Vulnerability Index (IDVI) that was introduced as a synthetic metric to account for a broader set of factors, including descriptors of health care, public health, economic, demographic, disease dynamics, and political (domestic and international) conditions [17]. While the SPAR indicator is a self-evaluation of countries capacity focusing on public health, the IDVI indicator includes broader sets of conditions that may have an impact on the management of a disease emergency.

Both SPAR and IDVI indicators range from 0 to 100, with increasing levels of capacity and decreasing vulnerability.

## Results

Egypt, Algeria and South Africa were the top three countries at highest introduction risk, with moderate to high SPAR capacity scores (87, 76 and 62, respectively) and IDVI (53, 49, 69) (Figure 2). Following countries in the introduction risk ranking include Nigeria and Ethiopia with moderate capacity (51, 67, respectively), but high vulnerability (27, 38) and substantially larger populations potentially exposed (Figure 3). Morocco, Sudan, Angola, Tanzania, Ghana and Kenya have similar moderate importation risk and population sizes. However, these countries present variable levels of capacity (ranging from 34 to 75) and an overall low IDVI (<46) reflecting a high vulnerability (except Morocco, with IDVI equal to 56). All other countries had low to moderate importation risk and low to moderate IDVI, with a majority having relatively low SPAR capacity score, with the exception of Tunisia and Rwanda.

**Figure 2.**
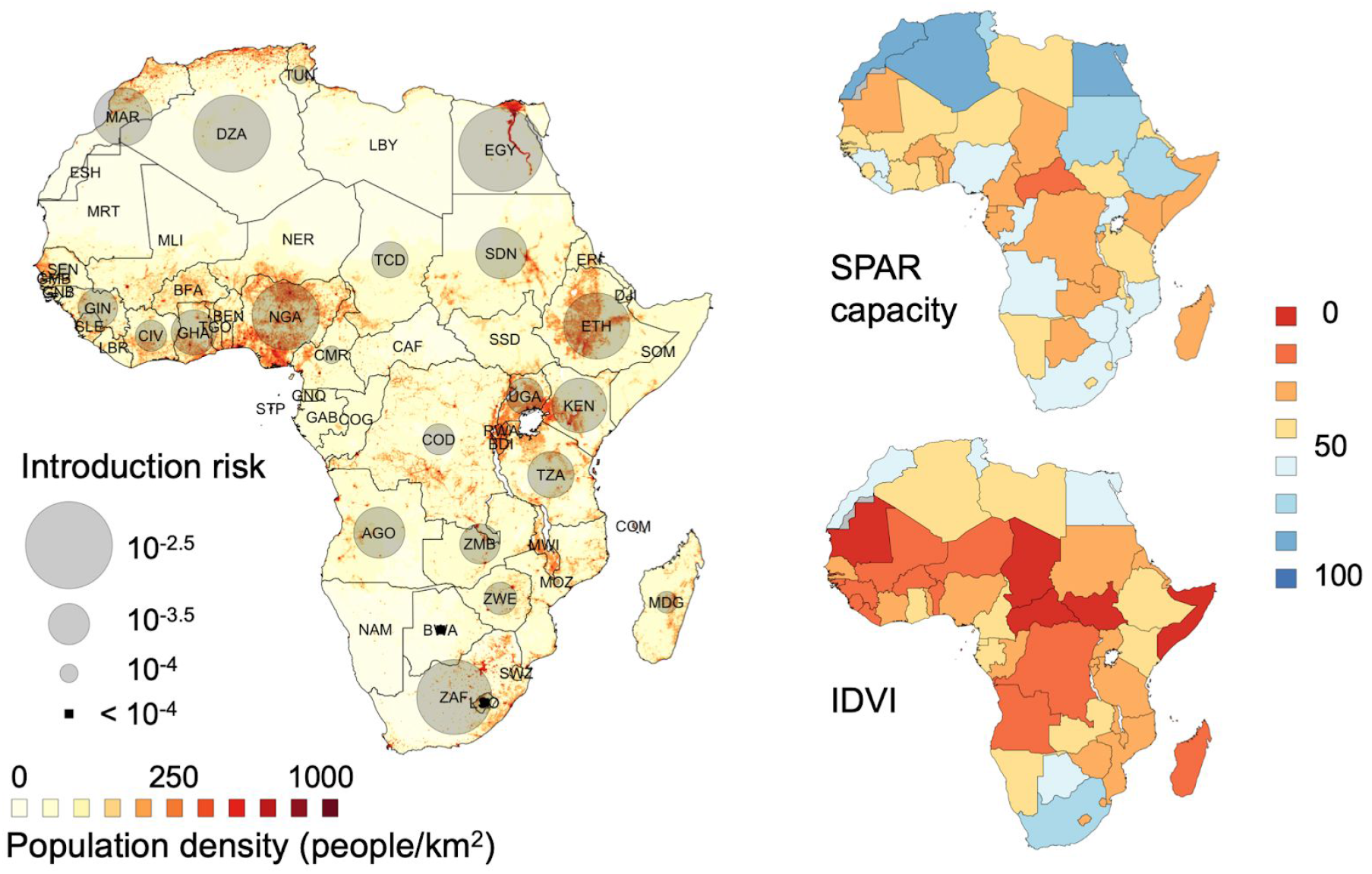
Global distribution of introduction risk over human population density (left) and distribution of the SPAR capacity index (top right) and Infectious Disease Vulnerability Index (IDVI, bottom right). Countries with no estimates of introduction risk correspond to situations where the risk of entry was found to be negligible at the time of analysis.

**Figure 3.**
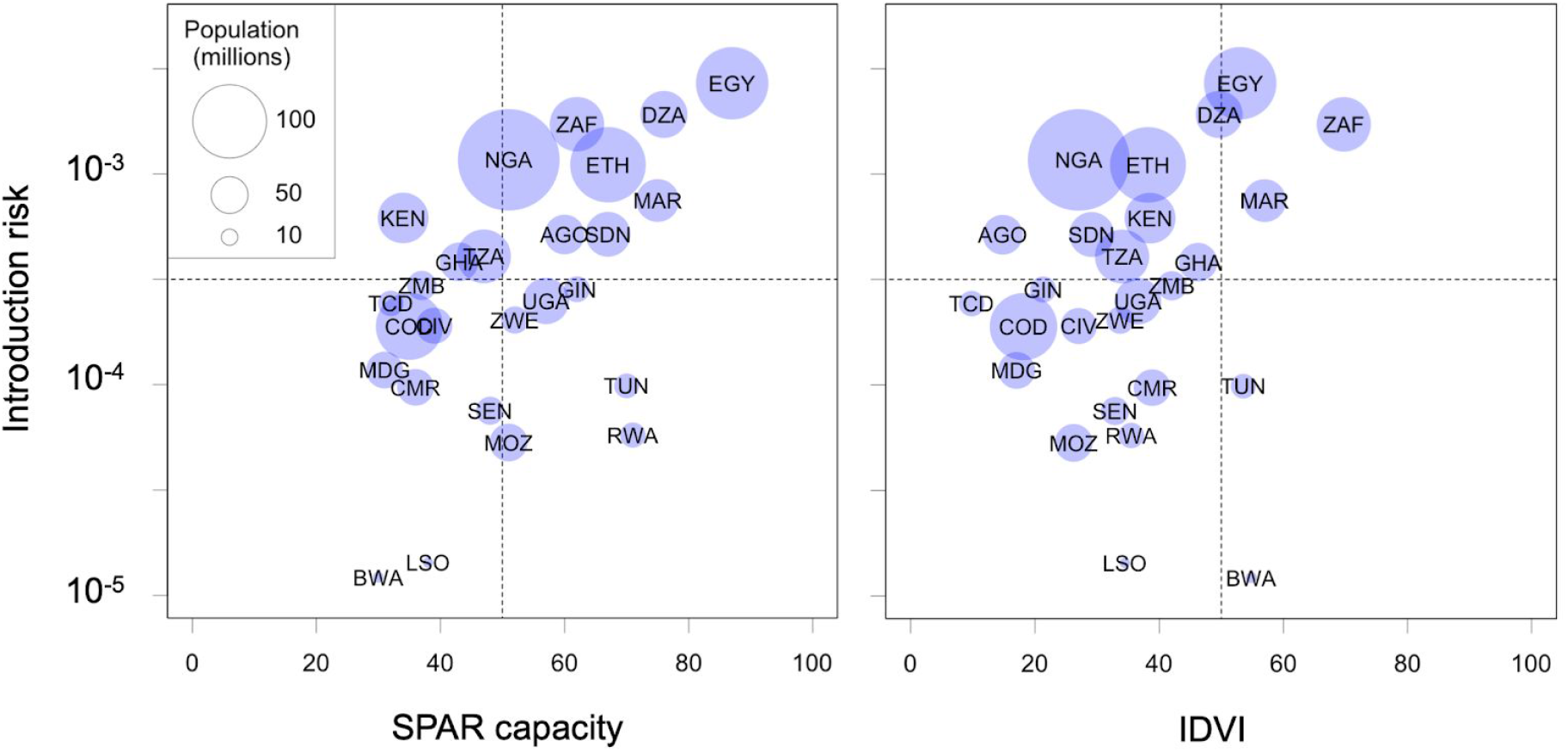
Introduction risk as a function of the SPAR capacity index (left) and Infectious Disease Vulnerability Index (IDVI, right) in Africa. Area of circles is proportional to country population.

Three clusters were identified among the countries with non-negligible risk (Figure 4). Each of the clusters corresponds to different Chinese airports as main source of entry risk (pie charts of Figure 4). The red cluster is highly exposed to Beijing province, moderately exposed to Guangdong and Shanghai provinces. The green cluster (including Botswana and Lesotho only) is exposed exclusively to the potential risk from airports in the Fujian province, and the blue cluster is heavily exposed to risk from Guangdong province and weakly to Zhejiang province.

**Figure 4.**
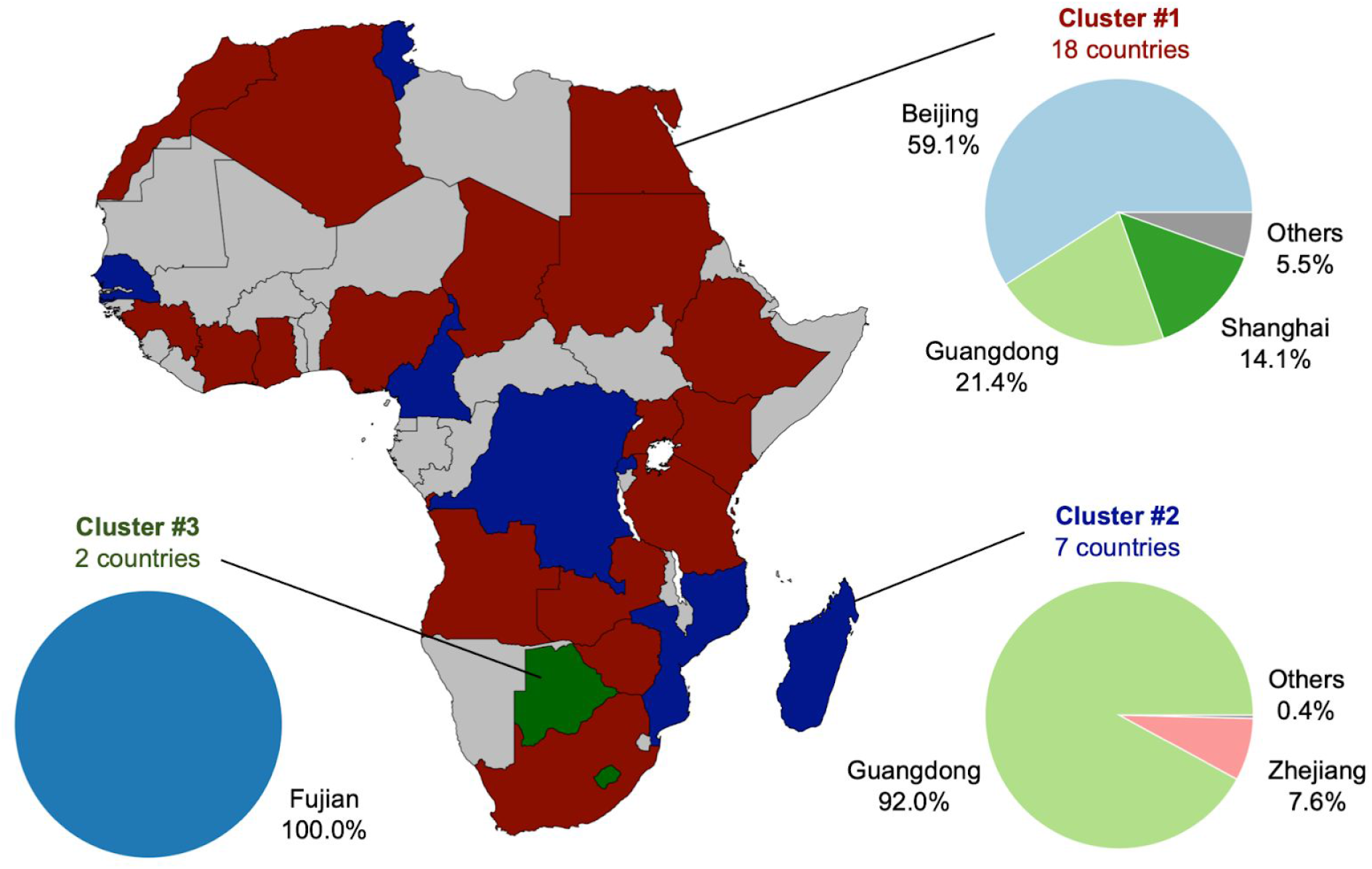
Cluster of countries sharing similar risk of importation from specific Chinese provinces (pie charts). Cluster #1 (red): Algeria, Angola, Chad, Egypt, Ethiopia, Ghana, Guinea, Ivory Coast, Kenya, Morocco, Nigeria, South Africa, Sudan, Tanzania, Uganda, Zambia, Zimbabwe. Cluster #2 (blue): Cameroon, Democratic Republic of the Congo, MadagascarMozambique, Rwanda, Senegal andTunisia. Cluster #3 (green): Botswana and Lesotho. Countries in grey were estimated to have a negligible risk of entry at the time of analysis.

## Discussion

Early detection of 2019-nCoV coronavirus importation and prevention of onward transmission are crucial challenges to all countries at risk of introduction from areas with active transmission in China. Seven countries in Asia, Europe, and North America have already reported secondary spread following importation. Secondary spread potentially occurring in countries with weaker health systems represents a major public health concern.

Here we show that the risk of introduction to African countries is highly heterogeneous, with Egypt, Algeria, South Africa, Ethiopia and Nigeria estimated to be at highest risk. We also identified that part of this heterogeneity in Africa depended on the distribution of cases within Chinese provinces. Shifts in local and widespread transmission in Beijing, Guangdong and Fujiian provinces could have profound implications for risk in Africa. For example, a significantly higher incidence in Guangdong than in other provinces would have a stronger impact on the importation risk of countries of cluster #2 than in countries from the other clusters. Also, flight bans put in place by some African airlines’ companies serving China [18] may alter the risk in the future. However, the main transporters continue to fly from China to Africa via their hubs in Africa or Middle East (e.g. Ethiopian Airlines, the largest carrier in Africa, today operating almost half of the flights from Africa to China [19]). While certain provinces contribute the largest to the risk of specific clusters of countries, enhanced surveillance at the airports should consider that importation may however still occur from those Chinese provinces that appear to have a lower probability in our estimations.

Algeria, Ethiopia, South Africa, Nigeria were part of the 13 top priority countries identified by WHO, based on their direct links and volume of travel to China [4]. Egypt, which was estimated here to be at highest risk, was not part of that list, although Cairo was identified as the African airport with the highest passenger volume from the affected areas [9]. Few other discrepancies were observed (Morocco and Angola were estimated to be at moderate risk but did not appear in the 13 top priority list) that may be explained by different risk estimation approaches. In our assessment, we accounted for the distribution of incidence within China and the volume of travel from China with the passenger network. This strongly impacts the spatial pattern in the risk of introduction. Yet, our data does not allow distinguishing between travel for tourism or business. Contrary to Europe, where the majority of cases so far were Chinese tourists, it may be expected that cases in Africa will be more business related, given the strong commercial links between African countries and China leading to frequent trips between the two continents.

Countries at the highest risk of importation, based on current epidemic situation in China, had moderate to high capacity scores. These may however correspond to different contributions to the mean SPAR indicators, reflecting different aspects of a country’s functional capacity. For example, South Africa had the maximum score for the laboratory capacity (100), but a low score in risk communication (20). Conversely, Nigeria had a fairly low score in the laboratory capacity (27) and the maximum score in the IHR coordination capacity (100). Conversely, countries with the lowest SPAR capacity score had moderate (Kenya, Tanzania, Ghana) to low introduction risk. The evaluation of additional factors (demographic, socio-economic, political) included in the IDVI that may influence the overall potential impact of an unfolding epidemic identifies several countries that had a significant importation risk with a low to medium IDVI, such as Nigeria, Ethiopia, Egypt and Algeria. The risk of importation from other points of entry, such as for example sea ports was not evaluated here.

Our results should be interpreted in relative terms. The overall SPAR score and IDVI of African countries is linked to their overall wealth, and are generally significantly lower than in many high-income countries, where the overall resource for detection, prevention and control are generally higher (SPAR ranging from 51 to 99 with a mean = 84.2 and IDVI from 78 to 97, with a mean = 88.3 in OECD countries). Comparatively, China has a SPAR score of 93 and an IDVI of 63.

African countries have recently strengthened their preparedness against 2019-nCoV importations [3,4,20]. Many countries improved airport surveillance and implemented temperature screening at ports of entry, including high-risk countries according to our analysis - South Africa [21], Ethiopia [22], and Nigeria [23], which also conducts interviews to passengers arriving from China. Overall recommendations to avoid travels to China have been issued. Communication campaigns have been intensified following WHO guidelines to provide information to health professionals and the general public, often with 24h dedicated hotlines, as in the case of Senegal [3].

Some countries remain however ill-equipped. Some lack the diagnostic for rapid testing for the virus, and, if cases are imported, tests will need to be performed abroad. This may critically increase the delay from identification of suspect cases to their confirmation and isolation, with an impact on possible disease transmission. WHO is currently supporting countries to improve their diagnostic capacity, previously limited to only two referral laboratories in the African region [4], and now extended also to Nigeria [24]. Also, resources to set up quarantine rooms for suspected cases at airports and hospitals, or to trace contacts of confirmed cases, as recommended by WHO, may be scarce. Countries may not have the same capacity to manage repatriations of nationals from the province of Hubei in China, as done by resource-rich countries, because of limited resources for quarantine and isolation. The epidemic in China highlights the rapid saturation of the hospital capacity if the outbreak is not contained. Increasing the number of available beds and supplies in resource-limited countries is critical in preparation to possible local transmission following importation. Crisis management plans should be ready in each African country.

Our findings help informing urgent prioritization for intensified support for preparedness and response in specific countries in Africa found to be at high risk and with relatively low capacity to manage the health emergency.

## Data Availability

All data are from the public domain, with links indicating their availability

https://www.worldpop.org

https://github.com/beoutbreakprepared/nCoV2019

https://extranet.who.int/e-spar

## Acknowledgments

This study was partially supported by the ANR project DATAREDUX (ANR-19-CE46-0008-03) to VC; the EU grant MOOD (H2020-874850) to MG, CP, MK, VC; Branco Weiss Fellowship to MK. We thank Rajesh Sreedharan for kind inputs on the use of IHR data, and REACTing (https://reacting.inserm.fr/) for useful discussions.

## Appendix

### Risk of case importation

The risk of importation of 2019-nCoV cases in a country outside China, α, from a city in China *i*, is based on:

– the travel flux from *i, n*_*i*_;
– the cumulated incidence in *i, e*_*i*_ (assumed to be homogeneous within each province);
– the probability of traveling from *i* to α, conditioned on traveling internationally from *i, A*_*i*α_ (by construction, 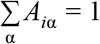).

We define risk flow from *i* to α as the matrix

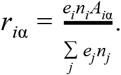

The risk of case importation to α from whatever origin in China is then

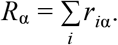

This risk is normalized so that 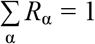

### Exposure analysis

For each African country, α, we define the exposure vector, *v*^(α)^, whose entry 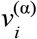 encodes the contribution of city *i* in China to the importation risk *R*_α_ :

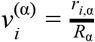

By construction these entries sum to one, 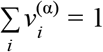. Therefore, we can use entropy-related metrics to quantify the similarity between the exposure patterns of two different destination countries, α and β. Specifically, once defined the entropy of *v*^(α)^ as

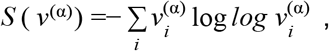

we used the Jensen-Shannon divergence between the two vectors, *v*^(α)^, and *v*^(β)^, defined as

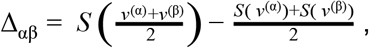

We then apply the complete linkage agglomerative clustering procedure to identify clusters of countries with similar exposure patterns.

